# BNT162b2 effectiveness against Delta & Omicron variants in teens by dosing interval and duration

**DOI:** 10.1101/2022.06.27.22276790

**Authors:** Iulia G. Ionescu, Danuta M. Skowronski, Chantal Sauvageau, Erica Chuang, Manale Ouakki, Shinhye Kim, Gaston De Serres

## Abstract

**Background and Objectives:** Two- and three-dose BNT162b2 (Pfizer-BioNTech) mRNA vaccine effectiveness (VE) against SARS-CoV-2 infection, including Delta and Omicron variants, was assessed among adolescents in two Canadian provinces where first and second doses were spaced longer than the manufacturer-specified 3-week interval.

**Methods:** Test-negative design estimated VE against laboratory-confirmed SARS-CoV-2 infection among 12-17-year-olds in Quebec and British Columbia, Canada between September 5, 2021 (epi-week 36), and April 30, 2022 (epi-week 17). Delta-dominant and Omicron-dominant periods spanned epi-weeks 36-47 and 51-17, respectively. VE was assessed from 14 days and explored by interval between first and second doses, time since second dose, and with administration of a third dose.

**Results:** Median first-second dosing-interval was ∼8 weeks and second-third dosing-interval was ∼29-31 weeks. Median follow-up post-second dose was ∼10-11 weeks for Delta-dominant and ∼21-22 weeks for Omicron-dominant periods, and ∼2-7 weeks post-third dose. VE against Delta was ≥90% to at least the 5th month post-second dose. VE against Omicron declined from ∼65-75% at 2-3 weeks to ≤50% by the 3^rd^ month post-vaccination, restored to ∼65% shortly following a third dose. VE exceeded 90% against Delta regardless of dosing-interval but appeared improved against Omicron with ≥8 weeks between first and second doses.

**Conclusion:** In adolescents, two BNT162b2 doses provided strong and sustained protection against Delta but reduced and rapidly-waning VE against Omicron. Longer interval between first and second doses and a third dose improved Omicron protection. Updated vaccine antigens, increased doses and/or dosing-intervals may be needed to improve adolescent VE against immunological-escape variants.

## Introduction

In May 2021, original authorization of the Pfizer-BioNTech BNT162b2 vaccine against COVID-19 for those ≥16 years old was extended in both the United States (US) and Canada to include adolescents 12-15 years of age.^1,2^ In phase III clinical trials, two-dose BNT162b2 efficacy against COVID-19 exceeded 90% in adolescents 7 days up to 4 months post-second dose.^1-3^ In Canada, the National Advisory Committee on Immunization (NACI) recommended a longer interval of up to four months between first and second doses of all COVID-19 vaccines beginning in March 2021. This extended dosing-interval was applied to adolescents when their vaccination began in two of the larger provinces of Canada, Quebec and British Columbia (BC).^4^ As vaccine supplies improved, both provinces shortened the dosing-interval to 8 weeks in late-May/early-June and to 4 weeks during the summer 2021 to increase vaccine coverage by autumn.^4^ By mid-September 2021, about two-thirds of 12-17-year-olds were twice-vaccinated (Supplementary Fig 1). Subsequently in October 2021, NACI recommended 8 weeks as the preferred dosing-interval, by which time about three quarters of adolescents in Quebec and BC were already twice-vaccinated, increasing to about 80% by mid-February 2022. Third dose administration for adolescents began in February 2022 in both provinces with an uptake of 14% and 34% respectively by end of April 2022. Considering the current context of the COVID-19 pandemic, third dose administration was recently recommended for all adolescents ≥6 and ≥5 months after the final dose in the primary COVID-19 vaccine series by the NACI and US Centers for Disease Control and Prevention (CDC), respectively.^5-6^

The Delta variant of concern (VOC) comprised virtually all SARS-CoV-2 detections in Quebec and BC between September and November 2021 before being displaced in December 2021 by Omicron, the latter almost sole-circulating thereafter through at least April 2022. In several post-marketing studies conducted elsewhere among adolescents, mRNA vaccine effectiveness (VE) exceeded 90% against infection and hospitalization during the pre-Omicron period.^7-16^ Fewer studies have assessed adolescent VE post-Omicron emergence,^14^ with the United Kingdom (UK) reporting rapid decline against Omicron beginning one month post-second dose.^13,17^ Here we report two-dose BNT162b2 VE among 12-17-year-olds in Quebec and BC, Canada between September 2021 and April 2022. We compare estimates during Delta-vs. Omicron-dominant periods and stratify by time since second dose, additionally capitalizing on unique immunization program adjustments to explore the effects of longer interval between first and second doses, as well as a third dose, in adolescents.

## Methods

### Study design, population and outcomes

A test-negative design (TND) was used to assess BNT162b2 VE against SARS-CoV-2 infection among adolescents. In Quebec VE was also assessed against symptomatic infection (not possible in BC). There are ∼500,000 12-17-year-olds in the province of Quebec and 300,000 in BC.^18^ In both provinces, specimens collected from community-dwelling adolescents between September 5, 2021 and April 30, 2022 and tested for SARS-CoV-2 by publicly-funded nucleic acid amplification test (NAAT) were eligible.

Logistic regression was used to estimate the adjusted odds ratio (AOR) for SARS-CoV-2 infection among two- and three-dose vaccinated compared to unvaccinated adolescents including as covariates: age group (12-14-year-olds and 15-17-year-olds), sex (female/male), calendar time (categorical by individual epi-week) and region. The latter included the 18 health administrative regions of Quebec, regrouped into five categories (Greater Montreal, Greater Quebec City, Central Quebec, Northern Quebec and others) and the five established health authorities in BC (Fraser, Interior, Vancouver Island, Northern and Vancouver Coastal). VE was derived as (1-AOR)x100%.

VE was assessed overall at ≥14 days post-vaccination and by time since second dose stratified by epidemic periods defined by VOC contribution. The Delta-dominant period, during which all cases were assumed to be due to Delta, spanned epi-weeks 36-47 (September 5 to November 27, 2021) when Delta comprised >99% of characterized viruses provincially.^19,20^ The period spanning epi-weeks 48-50 (November 28 to December 18, 2021) was considered a Delta-to-Omicron transition period. Finally, epi-weeks 51-17 (December 19, 2021 to April 30, 2022) were considered the Omicron-dominant period across which Omicron comprised >90% of characterized viruses and all cases were assumed to be due to Omicron in both provinces. VE was also explored by interval between first and second doses and with administration of a third dose.

All analyses were generated using SAS/STAT® software, Version 9.4 for Windows.

### Exclusion criteria

Participants identified as having had a NAAT-confirmed SARS-CoV-2 infection prior to September 5, 2021 were excluded as were those with missing information, vaccination that was invalid or involved <2 doses, two doses <21 days apart, three doses <24 weeks after the second, or with any vaccine other than BNT162b2.

### Cases and controls

Cases were 12-17-year-olds with NAAT-confirmed SARS-CoV-2 infection during the study period; controls were NAAT-negative. Individuals could contribute one test-positive specimen and were censored thereafter from case or control contribution. All test-negative specimens meeting inclusion criteria were used.

### Vaccination status

Participants were unvaccinated if they received no dose on or before specimen collection. VE was defined by second or third BNT162b2 receipt at least 14 days before the specimen collection date, allowing additional lag (compared to onset date) to mount an effective vaccine response, but a range of time since second dose was explored.

### Data sources

Specimens were sampled from provincial laboratory databases that captured all publicly-funded NAAT-testing provincially. In BC, additional case details were extracted through linkage with the provincial notifiable disease database capturing all SARS-CoV-2 cases reported to public health, including hospitalization information, the latter gathered in Quebec from administrative databases. Vaccination details were obtained through linkage with respective provincial registries. In Quebec, laboratory testing data also included testing indications at the time of sampling. Tests among symptomatic community-dwelling individuals had an “M7” specification as testing indication. Unique personal identifiers, distinct to each province, were used to perform individual-level database linkages for all participants.

### Ethics

Data linkages and analyses were authorized by the Provincial Health Officer (BC) and National Director of Public Health (Quebec) under respective provincial public health legislation without requirement for research ethics board review.

## Results

### Population profiles

In Quebec and BC, the study sample included 193,899 and 60,903 tests, respectively, of which 20,570 (11%) and 6,673 (11%), respectively, were test-positive cases, including 159 (0.8%) and 59 (0.9%), respectively, who were hospitalized (Table 1). Percent positivity among included specimens overall was consistent with the general population of adolescents (not shown), generally lower among the vaccinated versus unvaccinated, as expected (Supplementary Fig 1). No adolescent deaths were reported during the study period. Age and sex distributions were generally similar for cases and controls. In Quebec, cases were more often older adolescents and females than were controls and 54% of cases were symptomatic at specimen collection. Most cases occurred during the Omicron-dominant period, including 78% in Quebec and 57% in BC. In both provinces, median follow-up post-second dose was shorter during the Delta-(∼10-11 weeks) versus Omicron-dominant (∼21-22 weeks) period (Table 1).

**Table 1.** Profile of Participants (Adolescents Aged 12-17 Years), by Case/Control and Vaccine Status (Regardless of Time Since Vaccination), Quebec and British Columbia, Canada

| Characteristic | Quebec |  |  |  | British Columbia |  |  |  |
| --- | --- | --- | --- | --- | --- | --- | --- | --- |
|  | All (n=193899) |  | Two-dose vaccinated (n=169958) |  | All (n=60903) |  | Two-dose vaccinated (n=47831) |  |
|  | Cases, No.<br>(column %)<br>(n=20570) | Controls, No.<br>(column %)<br>(n=173329) | Cases, No.<br>(row %)<br>(n=16324) | Controls, No.<br>(row %)<br>(n=153634) | Cases, No.<br>(column %)<br>(n=6673) | Controls, No.<br>(column %)<br>(n=54230) | Cases, No.<br>(row %)<br>(n=3668) | Controls, No.<br>(row %)<br>(n=44163) |
| **Age, years** |  |  |  |  |  |  |  |  |
| 12-14 | 9476 (46.1) | 91942 (53.0) | 6780 (71.5) | 79087 (86.0) | 3392(50.8) | 27167 (50.1) | 1519 (44.9) | 20308 (74.8) |
| 15-17 | 11094 (53.9) | 81387 (47.0) | 9544 (86.0) | 74547 (91.6) | 3281 (49.2) | 27063 (49.9) | 2149 (65.5) | 23855 (88.1) |
| Median (IQR), years | 15 (13-16) | 14 (13-16) | 15 (13-16) | 14 (13-16) | 14 (13-16) | 14 (13-16) | 15 (13.5-16) | 15 (13-16) |
| **Sex** |  |  |  |  |  |  |  |  |
| Female | 11059 (53.8) | 88527 (51.1) | 9063 (82.0) | 78973 (89.2) | 3332 (49.9) | 26285 (48.5) | 1934 (58.0) | 21509 (81.8) |
| Male | 9511 (46.2) | 84802 (48.9) | 7261 (76.3) | 74661 (88.0) | 3341 (50.1) | 27945 (51.5) | 1734 (51.9) | 22654 (81.1) |
| **Infection severity** |  |  |  |  |  |  |  |  |
| Symptomatic | 11118 (54.0) | NA | 8912 (80.2) | NA | NA | NA | NA | NA |
| Hospitalization <sup>a</sup> | 159 (0.8) | NA | 123 (77.4) | NA | 59 (0.9) | NA | 33 (55.9) | NA |
| **Epidemiological period** |  |  |  |  |  |  |  |  |
| 36-47 (Sep 5, 2021 – Nov 27, 2021) | 2163 (10.5) | 106369 (61.4) | 514 (23.8) | 92458 (86.9) | 2437 (36.5) | 40660 (75.0) | 361 (14.8) | 32566 (80.1) |
| 48-50 (Nov 28, 2021 – Dec 18, 2021) | 2344 (11.4) | 35105 (20.3) | 1499 (64.0) | 31461 (89.6) | 448 (6.7) | 6415 (11.8) | 184 (41.1) | 5386 (84.0) |
| 51-17 (Dec 19,2021 – Apr 30, 2022) | 16063 (78.1) | 31855 (18.4) | 14311 (89.1) | 29715 (93.3) | 3788 (56.8) | 7155 (13.2) | 3123 (82.4) | 6211 (86.8) |
| **Time since second dose vaccination**<br>**(days/weeks)** | (all % values below this row are column %) |  |  |  |  |  |  |  |
| 0-13d (0-1wk) | NA | NA | 99 (0.6) | 2659 (1.7) | NA | NA | 45 (1.2) | 783 (1.8) |
| 14-27d (2-3wk) | NA | NA | 71 (0.4) | 5658 (3.7) | NA | NA | 21 (0.6) | 1485 (3.4) |
| 28-55d (4-7wk) | NA | NA | 289 (1.8) | 25113 (16.3) | NA | NA | 99 (2.7) | 7970 (18.0) |
| 56-83d (8-11wk) | NA | NA | 571 (3.5) | 33724 (22.0) | NA | NA | 223 (6.1) | 13990 (31.7) |
| 84-111d (12-15wk) | NA | NA | 1089 (6.7) | 28511 (18.6) | NA | NA | 290 (7.9) | 8357 (18.9) |
| 112-139d (16-19wk) | NA | NA | 3858 (23.6) | 29612 (19.3) | NA | NA | 514 (14.0) | 5918 (13.4) |
| 140-167d (20-23wk) | NA | NA | 6434 (39.4) | 18610 (12.1) | NA | NA | 1529 (41.7) | 4092 (9.3) |
| 168-195d (24-27wk) | NA | NA | 2224 (13.6) | 4347 (2.8) | NA | NA | 723 (19.7) | 977 (2.2) |
| 196-223d (28-31wk) | NA | NA | 910 (5.6) | 2366 (1.5) | NA | NA | 172 (4.7) | 329 (0.7) |
| 224-251d (32-35wk) | NA | NA | 524 (3.2) | 1838 (1.2) | NA | NA | 31 (0.8) | 153 (0.4) |
| 252-279d (36-39wk) | NA | NA | 242 (1.5) | 1050 (0.7) | NA | NA | 17 (0.5) | 100 (0.2) |
| 280-307d (40-43wk) | NA | NA | 9 (0.1) | 135 (0.1) | NA | NA | 3 (0.1) | 8 (0.0) |
| 308+d (44+wk) | NA | NA | 4 (0.0) | 11 (0.0) | NA | NA | 1 (0.0) | 1 (0.0) |
| Median (IQR), days | NA | NA | 149 (130-167) | 93 (60-130) | NA | NA | 153 (129-168) | 78 (57-114) |
| Median (IQR), days (epi-weeks 36-47) | NA | NA | 83 (55-107) | 68 (48-91) | NA | NA | 77 (51-103) | 68 (52-87) |
| Median (IQR), days (epi-weeks 48-50) | NA | NA | 133 (118-146) | 126 (110-139) | NA | NA | 130 (118-143) | 128 (112-139) |
| Median (IQR), days (epi-weeks 51-17) | NA | NA | 153 (135-170) | 153 (133-178) | NA | NA | 157 (142-171) | 151 (135-167) |
| **Interval between 1st and 2nd doses**<br>**(days/weeks)** |  |  |  |  |  |  |  |  |
| 21-34d (3-4wk) | NA | NA | 2510 (15.4) | 20669 (13.5) | NA | NA | 331 (9.0) | 2973 (6.7) |
| 35-48d (5-6wk) | NA | NA | 3803 (23.3) | 36195 (23.6) | NA | NA | 507 (13.8) | 4976 (11.3) |
| 49-62d (7-8wk) | NA | NA | 5144 (31.5) | 50293 (32.7) | NA | NA | 2234 (60.9) | 28715 (65.0) |
| 63-83d (9-11wk) | NA | NA | 4198 (25.7) | 40424 (26.3) | NA | NA | 505 (13.8) | 6459 (14.6) |
| 84-111d (12-15wk) | NA | NA | 538 (3.3) | 4916 (3.2) | NA | NA | 67 (1.8) | 829 (1.9) |
| 112+d (16+wk) | NA | NA | 131 (0.8) | 1137 (0.7) | NA | NA | 24 (0.7) | 211 (0.5) |
| Median (IQR), days | NA | NA | 54 (42-66) | 55 (43-66) | NA | NA | 54 (49-59) | 55 (50-60) |

Abbreviation: d, days; IQR, interquartile range; NA, not applicable; wk, weeks.
<sup>a</sup> COVID-19 related hospitalization occurring within 30 days of the reference date.

### Vaccination profiles

Cases and controls had similar median intervals between first and second dose of ∼8 weeks (54 and 55 days, respectively) (Table 1). However, more in Quebec were re-vaccinated at <7-week interval between doses for both cases and controls (39% and 37%, respectively) compared to BC (23% and 18%, respectively) (Table 1). Overall, 63% of cases in Quebec and 68% in BC occurred ≥20 weeks post-second dose, mostly during the Omicron-dominant period.

### Vaccine effectiveness

#### By outcome

During the Delta-dominant period, adjusted two-dose VE against any SARS-CoV-2 infection ≥14 days post-second dose exceeded 95% in both provinces (Fig 1, Supplementary Table 1). During the Delta-to-Omicron transition period, VE was slightly lower at 82.8% (95%CI: 81.0-84.4) in Quebec and 88.0% (95%CI: 85.1-90.3) in BC. During the Omicron-dominant period, VE decreased further to 41.9% (95%CI: 37.7-45.8) and 33.9% (95%CI: 25.7-41.1), respectively.

**Figure 1.**
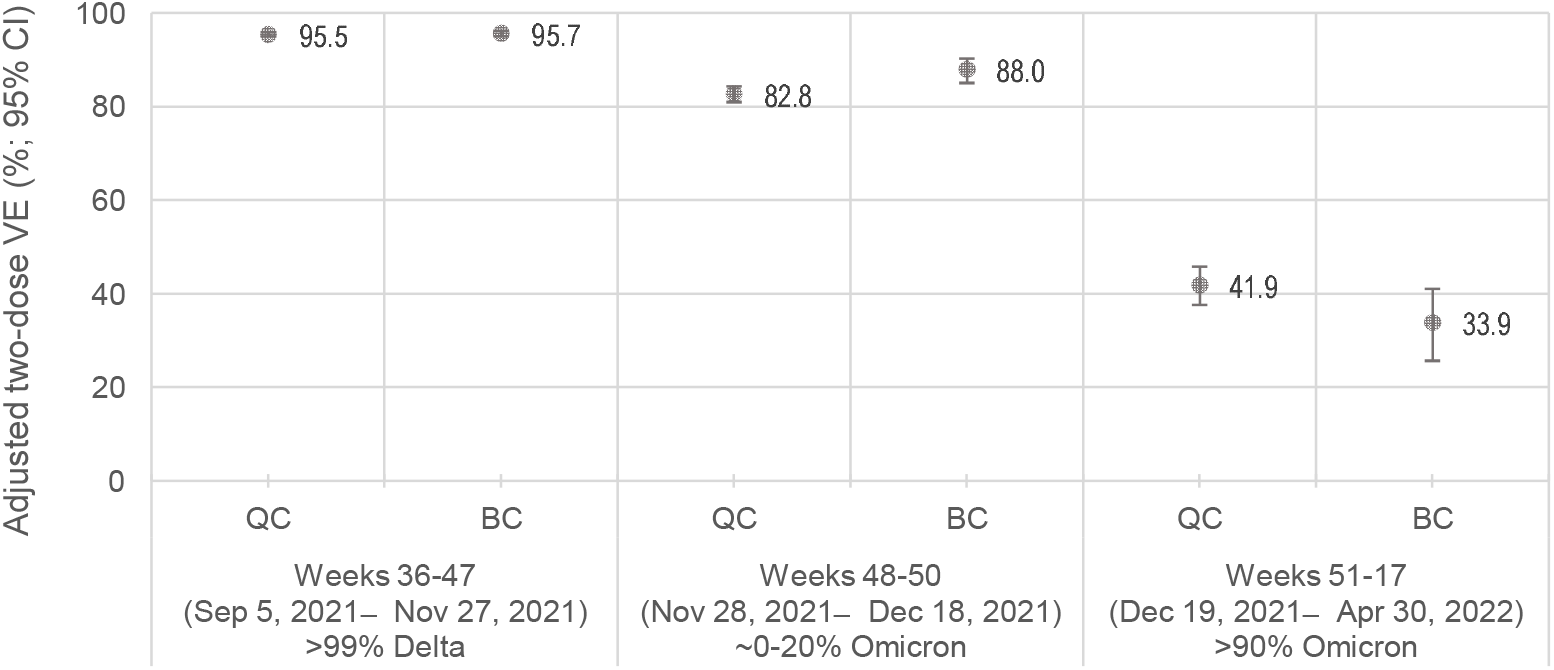
Adjusted Two-Dose BNT162b2 Vaccine Effectiveness Against Infection by Epidemiological Period, 12-17-Year-Olds, 14+ Days Post-Vaccination, Quebec and British Columbia, Canada Abbreviation: VE, vaccine effectiveness. Note: All estimates are adjusted for age, sex, epi-week and region.

In Quebec, adjusted two-dose VE was slightly higher against symptomatic vs. any SARS-CoV-2 infection during the Delta-dominant (97.3%; 95%CI: 96.8-97.7 vs. 95.5%; 95%CI: 95.0-96.0), and Delta-Omicron transition periods (87.9%; 95%CI: 86.1-89.5 vs. 82.8%; 95%CI: 81.0-84.4) and more so during the Omicron-dominant period (55.2%; 95%CI: 49.5-60.3 vs. 41.9%; 95%CI: 37.7-45.8) (Supplementary Fig 2, Supplementary Table 2).

#### By time since second dose

In both provinces, two-dose VE during the Delta-dominant period exceeded 95% from 2-3 weeks to at least the 3^rd^ month post-vaccination and remained >90% by the 5^th^ month post-vaccination (Fig 2, Supplementary Table 3). During the Delta-Omicron transition period, two-dose VE was >80% from 2–3 weeks post-vaccination and through at least the 5^th^ month. During the Omicron-dominant period, two-dose VE was lower overall, declining rapidly from ∼65-75% at 2-3 weeks post-vaccination to ∼40-50% by the 3^rd^ through 5^th^ months post-vaccination. During the 6^th^ month post-vaccination, two-dose VE against Omicron was 44.6% (95%CI: 40.0-49.0) in Quebec and 33.9% (95%CI: 24.1-42.4) in BC and by the 7^th^ month was 33.9% (95%CI: 27.4-39.9) and 22.2% (95%CI: 8.4-33.9), respectively.

**Figure 2.**
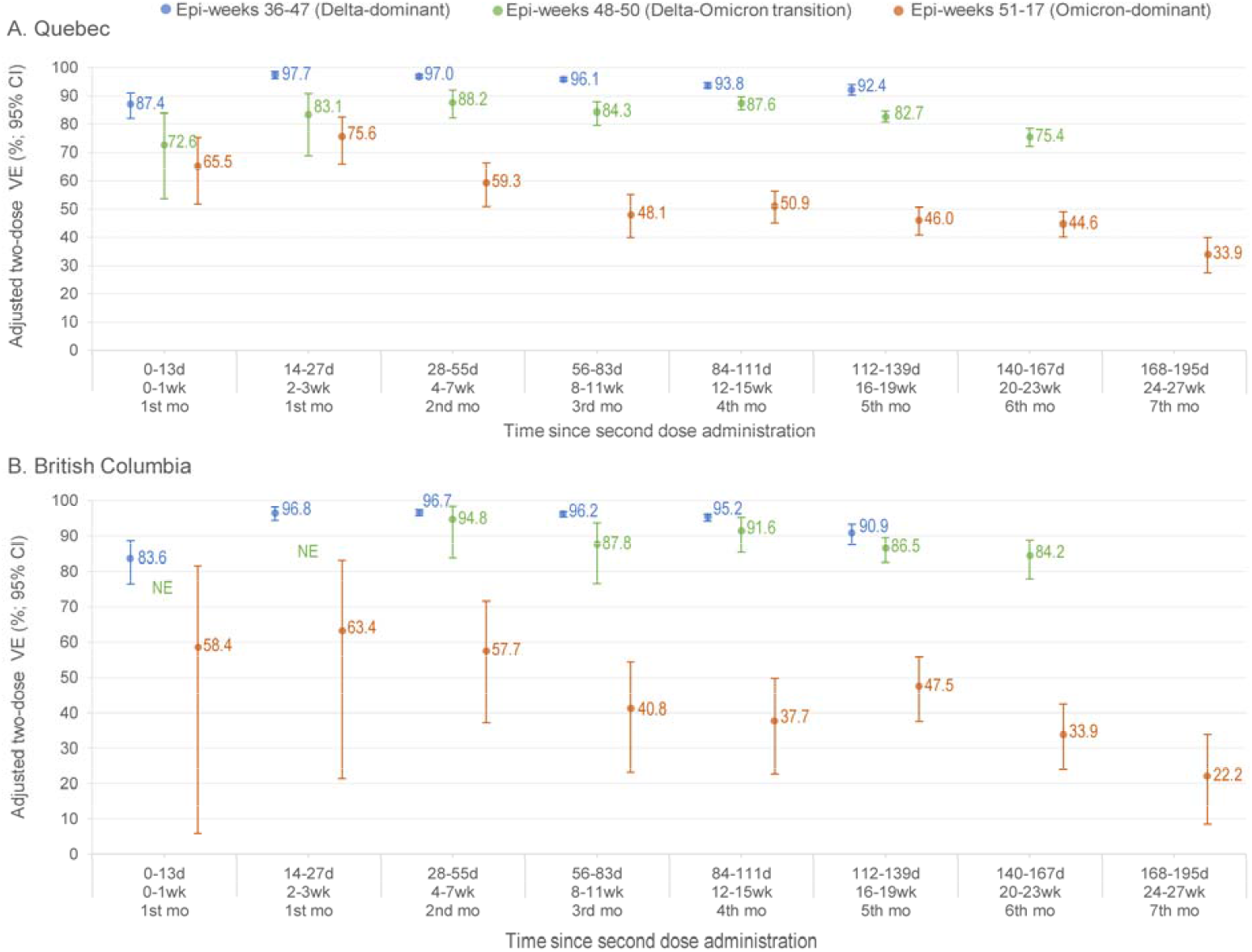
Adjusted Two-Dose BNT162b2 Vaccine Effectiveness Against Infection by Time Since Second Dose Administration and Epidemiological Period, 12-17-Year-Olds, Quebec (A) and British Columbia (B), Canada Abbreviation: d, days; mo, months; NE, not estimable or 95% confidence interval spans ≥100%; VE, vaccine effectiveness; wk, weeks. Note: All estimates are adjusted for age, sex, epi-week and region. Vaccine effectivenes estimates are displayed for a Delta-dominant period (blue), Delta-Omicron transition period (green) and Omicron-dominant period (red).

#### By interval between dose one and two

VE against Delta exceeded 90% in both provinces regardless of interval between first and second doses (Fig 3, Supplementary Table 4). Similarly, VE exceeded 80% regardless of dosing-interval during the Delta-Omicron transition period although variation in VOC contribution complicates that interpretation (Supplementary Table 4). Against Omicron, VE at 3-4-week vs. ≥8-week dosing-interval was 37.7% (95%CI: 32.1-42.8) vs. 44.5% (95%CI: 40.3-48.5), respectively, in Quebec and 30.0% (95%CI: 15.9-41.7) vs. 35.8% (95%CI: 26.9-43.7), respectively, in BC.

**Figure 3.**
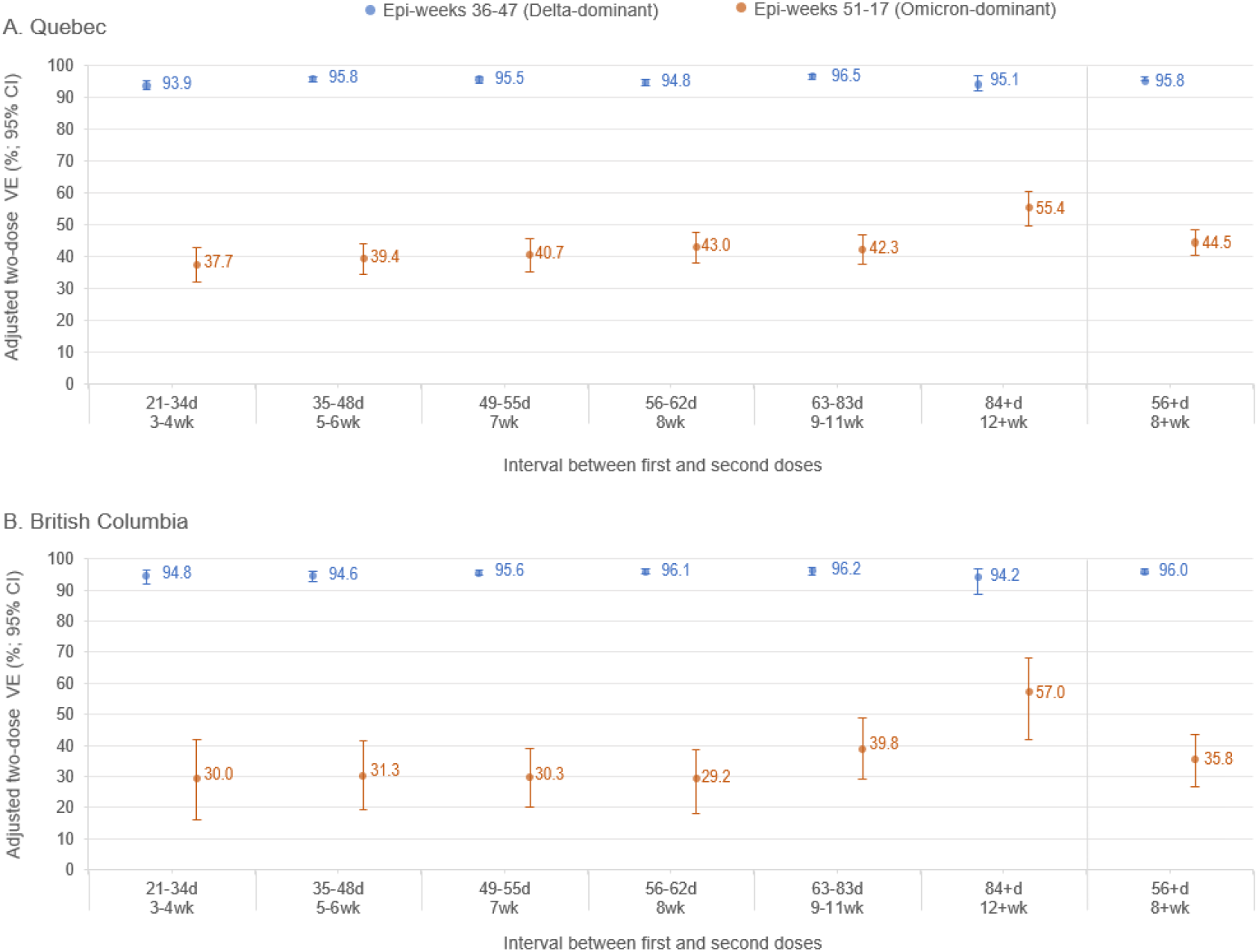
Adjusted Two-Dose BNT162b2 Vaccine Effectiveness Against Delta (Epi-Weeks 36-47) and Omicron (Epi-Weeks 51-17) Infection by Interval Between the First and Second Doses, 12-17-Year-Olds, 14+ Days Post-Vaccination, Quebec (A) and British Columbia (B), Canada Abbreviation: d, days; VE, vaccine effectiveness; wk, weeks. Note: All estimates are adjusted for age, sex, epi-week and region. Vaccine effectiveness estimates are displayed for a Delta-dominant period (blue) and Omicron-dominant period (red).

Improved Omicron VE with longer dosing-interval was most evident with ≥12-week interval between doses at 55.4% (95%CI: 49.6-60.5) in Quebec and 57.0% (95%CI: 42.0-68.1) in BC. However, when simultaneously stratifying for both dosing-interval and time since second dose (Supplementary Fig 3, Supplementary Table 5), VE against Omicron was not consistently higher for those vaccinated at ≥12-week vs. 8-11-week dosing-interval, with VE for both intervals typically exceeding that of shorter 5-7- and 3-4-week intervals at each monthly time point since second dose. A notable exception was the VE at three and four months post-second dose, highest among those vaccinated at ≥12-week interval in both provinces, but not consistently thereafter in either province. Reduced sample size was associated with wide confidence intervals, especially in BC.

#### Third dose

Median interval between second and third doses was ∼29-31 weeks with median follow-up post-third dose of ∼2-7 weeks (Supplementary Tables 6-7). Adjusted three-dose VE against Omicron at ≥14 days was 63.3% (95%CI: 47.2-74.6) in Quebec and 65.7% (95%CI: 41.9-79.8) in BC, recognizing lower third dose coverage and statistical power (Supplementary Tables 8-9).

## Discussion

This study found similar patterns of two-and three-dose BNT162b2 VE among 12-17-year-olds. Findings from both provinces suggest strong and sustained two-dose BNT162b2 protection against Delta variant infection, exceeding 90% to at least 5 months post-vaccination. Conversely, two-dose VE against the phylogenetically- and antigenically-distinct Omicron variant was reduced, rapidly declining from ∼65-75% at 2-3 weeks to ≤50% by 3 months post-vaccination. Whereas protection against Delta was high regardless of dosing-interval, VE against Omicron may have been improved by a longer (≥8 week) interval between first and second doses. Exploratory analysis suggests that shortly following a third dose, VE against Omicron was restored to the level observed 2-3 weeks after the second dose.

Our findings are consistent with efficacy data from the pivotal BNT162b2 clinical trial and results from observational studies that have evaluated BNT162b2 VE in adolescents prior to Omicron emergence. The phase III randomized trial with BNT162b2 mRNA vaccine in 12-15-year-olds reported vaccine efficacy of 100% (95%CI: 75.3-100) ≥7 days (and <4 months) post-second dose against non-Delta/non-Omicron variants.^3^ Two cohort studies from Israel evaluated two-dose effectiveness with 3-week spacing between doses against SARS-CoV-2 infection but with very short follow-up. The first study in 12-15-year-olds reported VE of 91.5% (95%CI: 88.2-93.9) between days 8-28 post-second dose.^7^ The second study in 12-18-year-olds reported VE on days 7-21 post-second dose of 90% (95%CI: 88-92) overall and 93% (95%CI: 88-97) against symptomatic infection.^8^ In the US, cohort analysis in 12-17-year-olds reported VE of 92% (95%CI: 79-97) against SARS-CoV-2 ≥14 days post-second dose (spaced as per manufacturer recommendation).^9^ In South Korea, a nationwide cohort study in high school seniors (16-18-year-olds) reported unadjusted two-dose VE at ≥14 days post-vaccination of 99% (95%CI: 99-100) against Delta.^10^ Each of the above studies, however, reflect pre-Omicron observations.

Evidence for BNT162b2 protection post-Omicron in adolescents is more limited. The PROTECT cohort study (US) reported adjusted VE 14-149 days post-second dose of 87% (95%CI: 49-97) against Delta and 59% (95%CI: 22-79) against Omicron infection in 12-15-year-olds.^11^ In Norway, where an extended interval of 8-12 weeks between doses was also used, population-based cohort study among 16-17-year-olds reported two-dose VE against Delta of 93% (95%CI: 90-95) at 35-62 days and 84% (95%CI: 76-89) at ≥63 days post-vaccination. VE against Omicron infection peaked at 53% (95%CI: 43-62) at 7-34 days and decreased to 23% (95%CI: 3-40) at ≥63 days post-vaccination.^12^ A TND conducted by Powell et al in the UK among 12-15-year-olds, where an extended 8-12-week dosing-interval was also used, found two-dose VE against symptomatic infection 7-13 days post-vaccination of 93% (95%CI: 82-98) during Delta and 83% (95%CI: 78-87) during Omicron periods.^13^ For 16-17-year-olds, two-dose VE at 14-34 days post-vaccination was 96% (95%CI: 95-97) and 71% (95%CI: 69-73) during Delta and Omicron periods, respectively.^13^ By 10 weeks post-vaccination, VE declined to 84% (95%CI: 72-91) against Delta but fell even lower to just 23% (95%CI: 15-30) against Omicron.^13^ In a TND conducted by Buchan et al among 12-17-year-olds from Ontario, Canada, two-dose VE against symptomatic Omicron infection was 51% (95%CI: 38-61) in the 7-59 days post-second dose and 29% (95%CI: 17-38) after 180 days.^14^ Two-dose VE against symptomatic Delta infection was 97% (95%CI: 94-99) and 90% (95%CI: 79-95), respectively.^14^ The similarity of findings between UK, Norway and Canada, where extended dosing-intervals were used, is reassuring. Here, we additionally show that a third dose spaced on average 7 months after the second restored Omicron protection to about the same levels observed several weeks post-second dose; however, longer third dose follow-up is needed as are additional evaluations of third dose protection elsewhere. We identified one third dose VE study in adolescents, a US TND showing similar third dose VE against symptomatic Omicron of 71.1% (95%CI: 65.5-75.7) at 2-6.5 weeks post-vaccination.^21^

SARS-CoV-2 infections among adolescents rarely progress to severe symptoms requiring hospitalization, causing less severe illness overall and far less deaths than in adults.^22^ We had few hospitalized cases (and no deaths) in this young population precluding precise or reliable VE estimation against those outcomes. Among 12-18-year-olds enrolled from 19 pediatric hospitals, two-dose BNT162b2 VE based on TND in the US was 93% against COVID-19 hospitalization during a period of Delta-dominance.^15^ An update with additional participants similarly showed VE of 94% (95%CI: 90-96) against Delta-associated hospitalization.^16^ Buchan et al estimated two-dose VE of 85% (95%CI: 74-91) against Omicron-associated hospitalizations ≥7 days post-vaccination in Ontario adolescents.^14^

The strengths of our study include large number of NAAT-confirmed SARS-CoV-2 cases, long follow-up after the second dose and estimates against both Delta and Omicron variants, standardized for time since second dose and interval between doses, as well as early exploration of third dose improvement. Alongside the UK and Norway, Canada is among few countries where VE based upon an extended interval between first and second doses can be assessed, including among adolescents. Third dose generalizability of our findings to other countries is possible where longer dosing-intervals were recommended between second and third doses, as is the case in the US. However, this study also has limitations. As for any observational study, residual confounding from unmeasured factors associated with both the likelihood of SARS-CoV-2 vaccination and infection cannot be ruled out. We did not have information allowing us to adjust for household size, younger siblings (ineligible for vaccination), or socio-economic status nor could we stratify estimates by underlying condition such as immune compromise. Data were sourced from surveillance registries, laboratory submissions and other administrative databases, subject to misclassification and selection bias. Changes in testing policies and behavioural factors can influence VE estimation. Differential healthcare-seeking behaviours may influence the probability of getting tested for SARS-CoV-2 because people who are vaccinated may also be more likely to seek medical care when ill.^23^ However, the TND partially reduces bias associated with this behaviour. Estimates were slightly higher (≤10%) in Quebec when analyses were restricted to testing for symptomatic versus any infection, reflecting the mostly symptom-based testing in both provinces. In both provinces, testing policies changed in mid-January to target mostly those at higher risk of severe outcomes or their caregivers, which may have influenced VE estimates against Omicron in an uncertain direction. Reduced sample sizes in further sub-stratified analyses by time since vaccination and interval between doses, and for third doses, affected statistical power and the precision of certain VE estimates. Vaccine passports or cards were required to attend social or recreational settings, applicable to adolescents in both provinces starting September 2021, and lifted in mid-March (Quebec) or early-April (BC). Although this might have led to greater exposure risk among vaccinated adolescents, potentially under-estimating VE, VE remained consistently high through November and the Delta-dominant period. Finally, our two-dose VE findings reflect a longer dosing-interval, as also applied in the UK and Norway, that require consideration in comparing to estimates elsewhere. Although a longer 8-week dosing-interval has recently been endorsed as the optimal interval between first and second mRNA doses by Canada’s NACI, as well as the World Health Organization and CDC, this may limit the generalizability of our findings to other areas using the 3-week manufacturer-specified schedule.^24-26^

## Conclusion

In conclusion, our findings indicate that two doses of BNT162b2 provided high protection against Delta infection in adolescents, maintained ≥90% up to 5 months post-second dose. Conversely, two-dose VE was more limited against Omicron infection, reducing the risk by up to three-quarters at 2-3 weeks post-vaccination, but by half or less within just a few months. Omicron VE may have been improved with longer interval between first and second doses, reinforcing some expert committee recommendations for preferred 8-week vs. manufacturer-specified 3-week interval between doses.^24-26^ Our findings are consistent with a greater antigenic distance between the vaccine strain and Omicron vs. Delta. Fortunately, hospitalization risk remains low among adolescents. However, if the goal of adolescent vaccination otherwise is to contribute to herd immunity, updated vaccine antigens, increased doses and/or dosing-intervals may be needed to improve adolescent VE against immunological-escape variants.

## Data Availability

Sharing of de-identified data that underpin the vaccine effectiveness results reported in this article will be considered upon request with appropriate review and aggregation as required to comply with respective provincial/national privacy and confidentiality legislation and through secure file transfer protocols. Information regarding submitting proposals and accessing data may be obtained through the corresponding authors.

## Abbreviations

AOR: adjusted odds ratio
CI: confidence interval
COVID-19: coronavirus disease 2019
mRNA: messenger RNA
NAAT: nucleic acid amplification test
NACI: Canadian National Advisory Committee on Immunization
SARS-CoV-2: severe acute respiratory syndrome coronavirus 2
TND: test-negative design
VE: vaccine effectiveness
VOC: variant of concern

## Acknowledgments

Authors in British Columbia thank Solmaz Setayeshgar for her earlier coding and analytical support and Hind Sbihi for her contributions in summarizing BC population laboratory findings, both of the BC Centre for Disease Control. Authors in both provinces thank the many frontline, regional and provincial practitioners, including clinical, laboratory and public health providers, epidemiologists, Medical Health Officers, laboratory staff, vaccinators, participants and others who contributed to the epidemiological, virological and genetic characterization data underpinning these analyses.

