## Supplementary Material for "BNT162b2 effectiveness against Delta & Omicron variants in teens by dosing interval and duration"

**Supplementary Figure 1.** % Test Positivity by Vaccine Status and Second Dose Vaccine Coverage, by Epi-Week, Adolescents Aged 12-17 Years, Quebec (A) and British Columbia (B), Canada

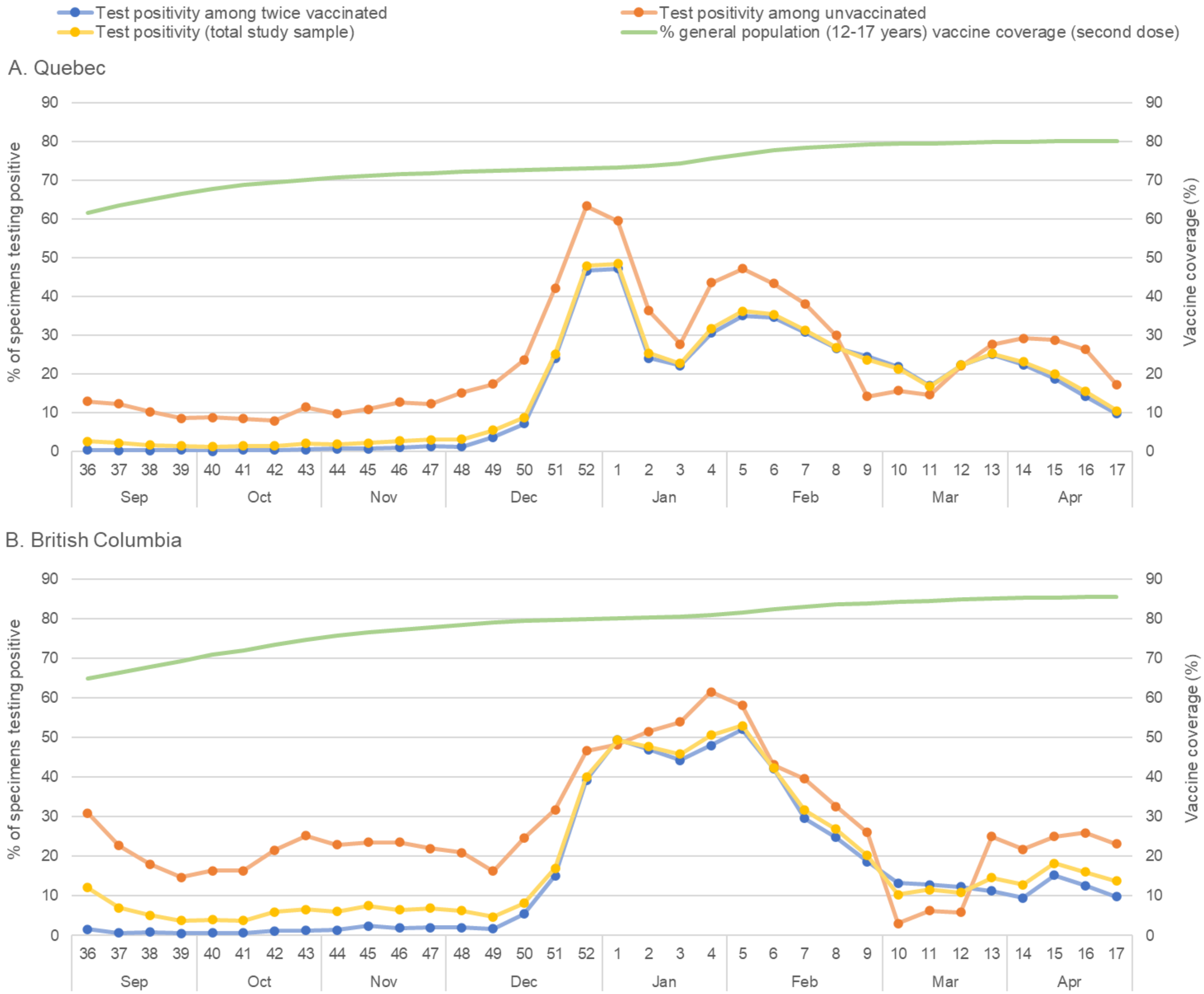

Note: A, Population vaccine coverage estimates based on vaccine information and population size by age group from the provincial immunization registry.[1] B, Population vaccine coverage estimates based on vaccine information from the provincial immunization registry with denominators based on estimates of population size by age group.[2]

[1] Reference: Données de vaccination contre la COVID-19 au Québec. INSPQ. Accessed March 3, 2022. Available from: https://www.inspq.qc.ca/covid-19/donnees/vaccination

[2] Reference for population size by age group: BC STATS. Population projections. BC Ministry of Citizens' Services, 2021. Accessed December 8, 2021. Available from: https://www2.gov.bc.ca/gov/content/data/statistics/people-population-community/population/population-projections

**Supplementary Table 1.** Unadjusted and Adjusted Two-Dose BNT162b2 Vaccine Effectiveness Against Infection by Vaccination Status and Epidemiological Period, 12-17-Year-Olds, Quebec and British Columbia, Canada

| Vaccination status | Quebec | | | | British Columbia | | | |
|---|---|---|---|---|---|---|---|---|
| | No. of cases | No. of controls | Unadjusted model VE (95% CI), % | Adjusted model [a] VE (95% CI), % | No. of cases | No. of controls | Unadjusted model VE (95% CI), % | Adjusted model [a] VE (95% CI), % |
| **Epi-weeks 36-47** | | | | | | | | |
| Unvaccinated | 1649 | 13911 | Reference | Reference | 2076 | 8094 | Reference | Reference |
| Two-dose vaccinated (0-13d) | 34 | 2261 | 87.3 (82.1, 91.0) | 87.4 (82.2, 91.0) | 31 | 721 | 83.2 (75.9, 88.3) | 83.6 (76.4, 88.6) |
| Two-dose vaccinated (14+d) | 480 | 90197 | 95.5 (95.0, 96.0) | 95.5 (95.0, 96.0) | 330 | 31845 | 96.0 (95.4, 96.4) | 95.7 (95.1, 96.2) |
| **Epi-weeks 48-50** | | | | | | | | |
| Unvaccinated | 845 | 3644 | Reference | Reference | 264 | 1029 | Reference | Reference |
| Two-dose vaccinated (0-13d) | 15 | 234 | 72.4 (53.2, 83.7) | 72.6 (53.6, 83.9) | 6 | 35 | NE | NE |
| Two-dose vaccinated (14+d) | 1484 | 31227 | 79.5 (77.6, 81.3) | 82.8 (81.0, 84.4) | 178 | 5351 | 87.0 (84.1, 89.4) | 88.0 (85.1, 90.3) |
| **Epi-weeks 51-17** | | | | | | | | |
| Unvaccinated | 1752 | 2140 | Reference | Reference | 665 | 944 | Reference | Reference |
| Two-dose vaccinated (0-13d) | 50 | 164 | 62.8 (48.6, 73.0) | 65.5 (51.7, 75.3) | 8 | 27 | 57.9 (6.8, 81.0) | 58.4 (5.9, 81.6) |
| Two-dose vaccinated (14+d) | 14261 | 29551 | 41.1 (37.0, 44.8) | 41.9 (37.7, 45.8) | 3115 | 6184 | 28.5 (20.3, 35.8) | 33.9 (25.7, 41.1) |

Abbreviation: d, days; NE, not estimable or 95% confidence interval spans ≥100%; VE, vaccine effectiveness.

[a] Adjusted for age, sex, epi-week and region.

**Supplementary Figure 2.** Adjusted Two-Dose BNT162b2 Vaccine Effectiveness Against All or Symptomatic Community-Based Infections by Epidemiological Period, 12-17-Year-Olds, 14+ Days Post-Vaccination, Quebec, Canada

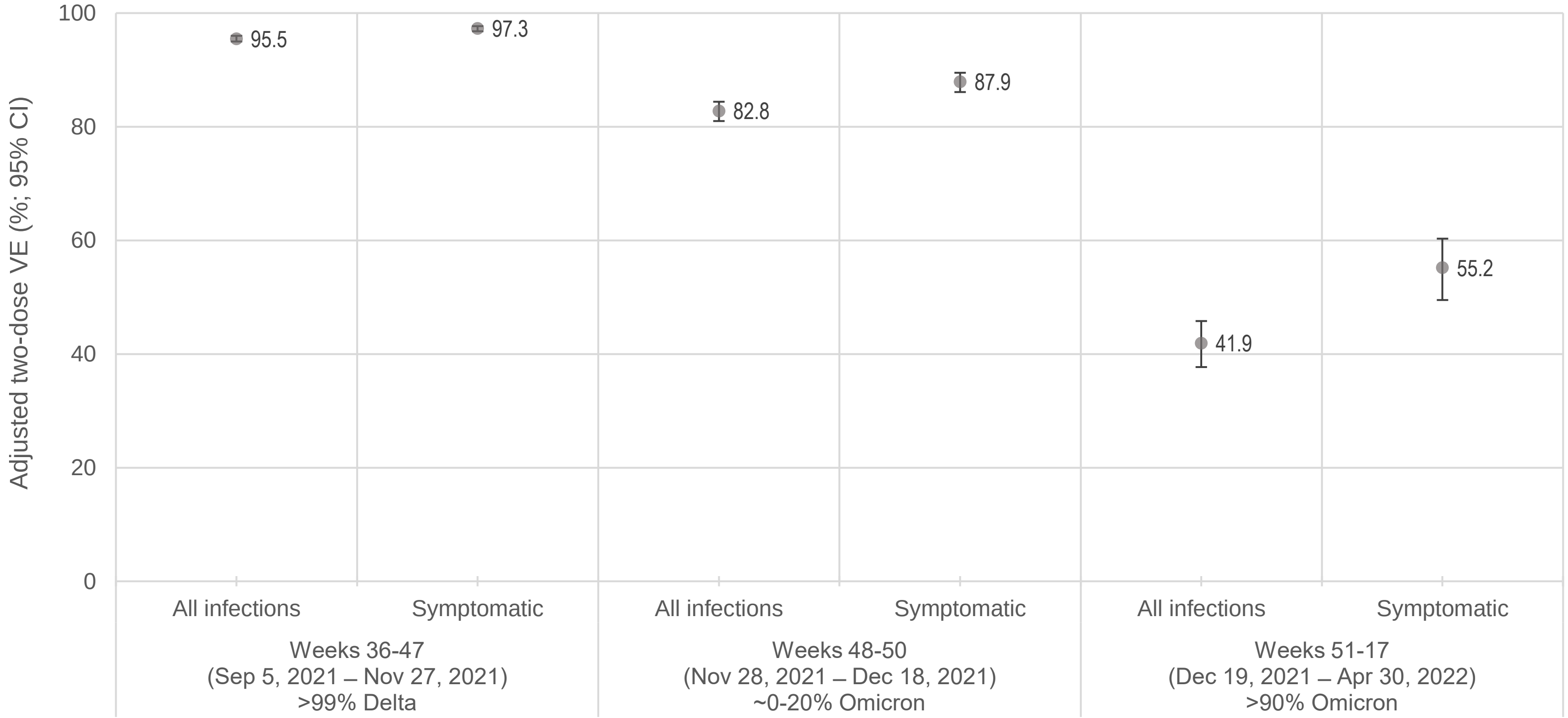

Abbreviation: VE, vaccine effectiveness.

Note: All estimates are adjusted for age, sex, epi-week and region. Estimates against symptomatic infection from Quebec are based on the "M7" code specified as testing indication on the laboratory requisition. Similar restriction based upon symptomatic presentation was not possible in British Columbia although testing in that province was foremost symptom-based overall.

**Supplementary Table 2.** Unadjusted and Adjusted Two-Dose BNT162b2 Vaccine Effectiveness Against Community-Based Symptomatic Infection by Vaccination Status and Epidemiological Period, 12-17-Year-Olds, Quebec, Canada

| | Quebec | | | |
|---|---|---|---|---|
| | | | Unadjusted model | Adjusted model [a] |
| **Vaccination status** | **No. of Cases** | **No. of controls** | **VE (95% CI), %** | **VE (95% CI), %** |
| **Epi-weeks 36-47** | | | | |
| Unvaccinated | 839 | 6855 | Reference | Reference |
| Two-dose vaccinated (0-13d) | 12 | 1168 | 91.6 (85.1, 95.3) | 91.7 (85.2, 95.3) |
| Two-dose vaccinated (14+d) | 193 | 57791 | 97.3 (96.8, 97.7) | 97.3 (96.8, 97.7) |
| **Epi-weeks 48-50** | | | | |
| Unvaccinated | 448 | 1140 | Reference | Reference |
| Two-dose vaccinated (0-13d) | 4 | 70 | 85.5 (59.9, 94.7) | 86.0 (61.1, 94.9) |
| Two-dose vaccinated (14+d) | 791 | 13246 | 84.8 (82.7, 86.7) | 87.9 (86.1, 89.5) |
| **Epi-weeks 51-17** | | | | |
| Unvaccinated | 919 | 466 | Reference | Reference |
| Two-dose vaccinated (0-13d) | 25 | 55 | 77.0 (62.5, 85.8) | 76.7 (61.3, 86.0) |
| Two-dose vaccinated (14+d) | 7887 | 9091 | 56.0 (50.6, 60.8) | 55.2 (49.5, 60.3) |

Abbreviation: d, days; VE, vaccine effectiveness.

[a] Adjusted for age, sex, epi-week and region.

**Supplementary Table 3.** Unadjusted and Adjusted Two-Dose BNT162b2 Vaccine Effectiveness Against Infection by Time Since Vaccination and Epidemiological Period, 12-17-Year-Olds, Quebec and British Columbia, Canada

| Time since 2nd dose vaccination | Quebec | | | | British Columbia | | | |
|---|---|---|---|---|---|---|---|---|
| | No. of cases | No. of controls | Unadjusted model VE (95% CI), % | Adjusted model [a] VE (95% CI), % | No. of cases | No. of controls | Unadjusted model VE (95% CI), % | Adjusted model [a] VE (95% CI), % |
| **Epi-weeks 36-47** | | | | | | | | |
| Unvaccinated | 1649 | 13911 | Reference | Reference | 2076 | 8094 | Reference | Reference |
| 0-13d (0-1wk; 1st mo) | 34 | 2261 | 87.3 (82.1, 91.0) | 87.4 (82.2, 91.0) | 31 | 721 | 83.2 (75.9, 88.3) | 83.6 (76.4, 88.6) |
| 14-27d (2-3wk; 1st mo) | 15 | 5146 | 97.5 (95.9, 98.5) | 97.7 (96.2, 98.6) | 12 | 1397 | 96.7 (94.1, 98.1) | 96.8 (94.4, 98.2) |
| 28-55d (4-7wk; 2nd mo) | 82 | 23655 | 97.1 (96.3, 97.7) | 97.0 (96.3, 97.6) | 60 | 7624 | 96.9 (96.0, 97.6) | 96.7 (95.7, 97.5) |
| 56-83d (8-11wk; 3rd mo) | 131 | 31159 | 96.5 (95.8, 97.0) | 96.1 (95.3, 96.7) | 105 | 13437 | 97.0 (96.3, 97.5) | 96.2 (95.3, 96.9) |
| 84-111d (12-15wk; 4th mo) | 162 | 21567 | 93.7 (92.5, 94.6) | 93.8 (92.7, 94.8) | 96 | 7316 | 94.9 (93.7, 95.8) | 95.2 (94.1, 96.2) |
| 112-139d (16-19wk; 5th mo) | 88 | 8326 | 91.1 (88.9, 92.8) | 92.4 (90.4, 94.0) | 53 | 1982 | 89.6 (86.2, 92.1) | 90.9 (87.7, 93.2) |
| **Epi-weeks 48-50** | | | | | | | | |
| Unvaccinated | 845 | 3644 | Reference | Reference | 264 | 1029 | Reference | Reference |
| 0-13d (0-1wk; 1st mo) | 15 | 234 | 72.4 (53.2, 83.7) | 72.6 (53.6, 83.9) | 6 | 35 | NE | NE |
| 14-27d (2-3wk; 1st mo) | 11 | 288 | 83.5 (69.8, 91.0) | 83.1 (68.9, 90.8) | 0 | 56 | NE | NE |
| 28-55d (4-7wk; 2nd mo) | 25 | 922 | 88.3 (82.5, 92.2) | 88.2 (82.3, 92.1) | 3 | 224 | 94.8 (83.6, 98.3) | 94.8 (83.7, 98.4) |
| 56-83d (8-11wk; 3rd mo) | 64 | 1754 | 84.3 (79.6, 87.9) | 84.3 (79.6, 87.9) | 10 | 319 | 87.8 (76.7, 93.6) | 87.8 (76.6, 93.6) |
| 84-111d (12-15wk; 4th mo) | 143 | 5331 | 88.4 (86.1, 90.4) | 87.6 (85.1, 89.7) | 14 | 670 | 91.9 (85.9, 95.3) | 91.6 (85.4, 95.2) |
| 112-139d (16-19wk; 5th mo) | 689 | 15225 | 80.5 (78.3, 82.5) | 82.7 (80.7, 84.6) | 97 | 2836 | 86.7 (83.0, 89.5) | 86.5 (82.5, 89.5) |
| 140-167d (20-23wk; 6th mo) | 535 | 7542 | 69.4 (65.7, 72.7) | 75.4 (72.1, 78.4) | 53 | 1211 | 82.9 (76.8, 87.4) | 84.2 (77.8, 88.8) |
| **Epi-weeks 51-17** | | | | | | | | |
| Unvaccinated | 1752 | 2140 | Reference | Reference | 665 | 944 | Reference | Reference |
| 0-13d (0-1wk; 1st mo) | 50 | 164 | 62.8 (48.6, 73.0) | 65.5 (51.7, 75.3) | 8 | 27 | 57.9 (6.8, 81.0) | 58.4 (5.9, 81.6) |
| 14-27d (2-3wk; 1st mo) | 45 | 224 | 75.5 (66.0, 82.3) | 75.6 (65.8, 82.6) | 9 | 32 | 60.1 (15.8, 81.1) | 63.4 (21.4, 83) |
| 28-55d (4-7wk; 2nd mo) | 182 | 536 | 58.5 (50.4, 65.3) | 59.3 (50.9, 66.3) | 36 | 122 | 58.1 (38.5, 71.5) | 57.7 (37.2, 71.6) |
| 56-83d (8-11wk; 3rd mo) | 376 | 811 | 43.4 (35.0, 50.6) | 48.1 (39.9, 55.1) | 108 | 234 | 34.5 (16.0, 48.9) | 40.8 (23.2, 54.4) |
| 84-111d (12-15wk; 4th mo) | 784 | 1613 | 40.6 (34.0, 46.6) | 50.9 (44.9, 56.3) | 180 | 371 | 31.1 (15.6, 43.8) | 37.7 (22.7, 49.7) |
| 112-139d (16-19wk; 5th mo) | 3081 | 6061 | 37.9 (33.0, 42.5) | 46.0 (40.9, 50.7) | 364 | 1100 | 53.0 (45.2, 59.7) | 47.5 (37.6, 55.8) |
| 140-167d (20-23wk; 6th mo) | 5897 | 10773 | 33.1 (28.2, 37.7) | 44.6 (40.0, 49.0) | 1472 | 2804 | 25.5 (16.2, 33.7) | 33.9 (24.1, 42.4) |
| 168-195d (24-27wk; 7th mo) | 2209 | 4176 | 35.4 (29.9, 40.4) | 33.9 (27.4, 39.9) | 722 | 944 | -8.6 (-24.7, 5.5) | 22.2 (8.4, 33.9) |

Abbreviation: d, days; mo, months; NE, not estimable or 95% confidence interval spans ≥100%; VE, vaccine effectiveness; wk, weeks.
[a] Adjusted for age, sex, epi-week and region.

**Supplementary Table 4.** Unadjusted and Adjusted Two-Dose BNT162b2 Vaccine Effectiveness Against Infection by Interval Between the First and Second Doses and Epidemiological Period, 12-17-Year-Olds, 14+ Days Post-Vaccination, Quebec and British Columbia, Canada

| Interval between the 1st and 2nd doses | Quebec | | | | British Columbia | | | |
|---|---|---|---|---|---|---|---|---|
| | No. of cases | No. of controls | Unadjusted model VE (95% CI), % | Adjusted model [a] VE (95% CI), % | No. of cases | No. of controls | Unadjusted model VE (95% CI), % | Adjusted model [a] VE (95% CI), % |
| **Epi-weeks 36-47** | | | | | | | | |
| Unvaccinated | 1649 | 13911 | Reference | Reference | 2076 | 8094 | Reference | Reference |
| 21-34d (3-4wk) | 77 | 10393 | 93.7 (92.1, 95.0) | 93.9 (92.3, 95.2) | 23 | 1785 | 95.0 (92.4, 96.7) | 94.8 (92.1, 96.6) |
| 35-48d (5-6wk) | 107 | 21837 | 95.9 (95.0, 96.6) | 95.8 (94.9, 96.6) | 44 | 3202 | 94.6 (92.8, 96.0) | 94.6 (92.7, 96.0) |
| 49-55d (7wk) | 75 | 14261 | 95.6 (94.4, 96.5) | 95.5 (94.4, 96.5) | 123 | 12384 | 96.1 (95.3, 96.8) | 95.6 (94.7, 96.4) |
| 56-62d (8wk) | 102 | 16553 | 94.8 (93.6, 95.7) | 94.8 (93.6, 95.7) | 87 | 9253 | 96.3 (95.4, 97.0) | 96.1 (95.1, 96.9) |
| 63-83d (9-11wk) | 103 | 24365 | 96.4 (95.6, 97.1) | 96.5 (95.7, 97.1) | 44 | 4658 | 96.3 (95.0, 97.3) | 96.2 (94.8, 97.2) |
| 84+d (12+wk) | 16 | 2788 | 95.2 (92.1, 97.0) | 95.1 (92.0, 97.0) | 9 | 563 | 93.8 (87.9, 96.8) | 94.2 (88.7, 97.0) |
| 56+d (8+wk) | 221 | 43706 | 95.7 (95.1, 96.3) | 95.8 (95.1, 96.3) | 140 | 14474 | 96.2 (95.5, 96.8) | 96.0 (95.3, 96.7) |
| **Epi-weeks 48-50** | | | | | | | | |
| Unvaccinated | 845 | 3644 | Reference | Reference | 264 | 1029 | Reference | Reference |
| 21-34d (3-4wk) | 218 | 4724 | 80.1 (76.8, 83.0) | 81.6 (78.4, 84.2) | 14 | 404 | 86.5 (76.6, 92.2) | 86.1 (75.8, 92.0) |
| 35-48d (5-6wk) | 365 | 7415 | 78.8 (75.8, 81.3) | 80.6 (77.8, 83.0) | 23 | 702 | 87.2 (80.2, 91.7) | 87.4 (80.4, 91.9) |
| 49-55d (7wk) | 225 | 4538 | 78.6 (75.1, 81.7) | 80.8 (77.4, 83.6) | 70 | 1894 | 85.6 (81.0, 89.0) | 85.4 (80.4, 89.1) |
| 56-62d (8wk) | 268 | 5387 | 78.5 (75.2, 81.4) | 80.4 (77.3, 83.1) | 43 | 1444 | 88.4 (83.8, 91.7) | 88.4 (83.6, 91.8) |
| 63-83d (9-11wk) | 373 | 8123 | 80.2 (77.5, 82.6) | 82.4 (79.9, 84.6) | 26 | 771 | 86.9 (80.1, 91.3) | 87.2 (80.4, 91.7) |
| 84+d (12+wk) | 35 | 1040 | 85.5 (79.5, 89.7) | 86.1 (80.3, 90.2) | 2 | 136 | 94.3 (76.7, 98.6) | 93.8 (74.4, 98.5) |
| 56+d (8+wk) | 676 | 14550 | 80.0 (77.7, 82.0) | 82.5 (80.4, 84.4) | 71 | 2351 | 88.2 (84.5, 91.0) | 88.7 (85.0, 91.5) |
| **Epi-weeks 51-17** | | | | | | | | |
| Unvaccinated | 1752 | 2140 | Reference | Reference | 665 | 944 | Reference | Reference |
| 21-34d (3-4wk) | 2191 | 4097 | 34.7 (29.1, 39.8) | 37.7 (32.1, 42.8) | 285 | 542 | 25.4 (11.1, 37.3) | 30.0 (15.9, 41.7) |
| 35-48d (5-6wk) | 3312 | 6661 | 39.3 (34.5, 43.7) | 39.4 (34.3, 44.2) | 424 | 832 | 27.7 (15.7, 37.9) | 31.3 (19.2, 41.6) |
| 49-55d (7wk) | 2088 | 4253 | 40.0 (34.9, 44.8) | 40.7 (35.3, 45.8) | 1056 | 2014 | 25.6 (15.7, 34.3) | 30.3 (20.0, 39.2) |
| 56-62d (8wk) | 2367 | 5075 | 43.0 (38.3, 47.4) | 43.0 (37.9, 47.7) | 848 | 1615 | 25.5 (15.2, 34.5) | 29.2 (18.3, 38.6) |
| 63-83d (9-11wk) | 3709 | 7714 | 41.3 (36.7, 45.5) | 42.3 (37.5, 46.7) | 431 | 954 | 35.9 (25.4, 44.9) | 39.8 (29.0, 49.0) |
| 84+d (12+wk) | 594 | 1751 | 58.6 (53.6, 63.0) | 55.4 (49.6, 60.5) | 71 | 227 | 55.6 (41.0, 66.6) | 57.0 (42.0, 68.1) |
| 56+d (8+wk) | 6670 | 14540 | 44.0 (39.9, 47.7) | 44.5 (40.3, 48.5) | 1350 | 2796 | 31.5 (22.8, 39.1) | 35.8 (26.9, 43.7) |

Abbreviation: d, days; VE, vaccine effectiveness, wk, weeks.
[a] Adjusted for age, sex, epi-week and region.

**Supplementary Figure 3.** Adjusted Two-Dose BNT162b2 Vaccine Effectiveness Against Omicron Infection (Epi-Weeks 51-17) by Time Since Second Dose Administration and Interval Between the First and Second Doses, 12-17-Year-Olds, Quebec (A) and British Columbia (B), Canada

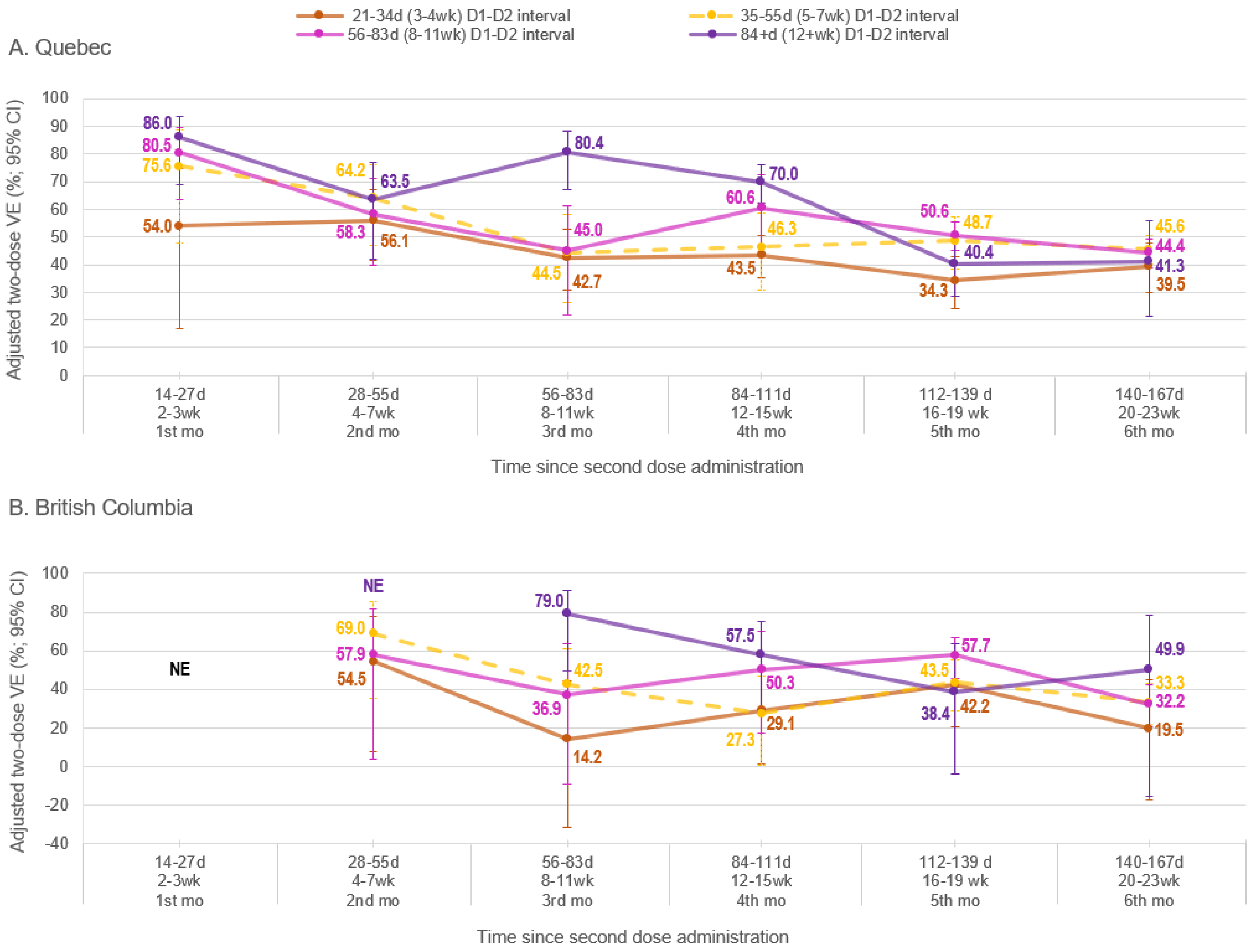

Abbreviation: d, days; D1, 1st dose; D2, 2nd dose; mo, months; NE, not estimable or 95% confidence interval spans ≥100%; VE, vaccine effectiveness ; wk, weeks.

Note: All estimates are adjusted for age, sex, epi-week and region.

**Supplementary Table 5.** Unadjusted and Adjusted Two-Dose BNT162b2 Vaccine Effectiveness Against Omicron Infection (Epi-Weeks 51-17) by Time Since Second Dose Administration and Interval Between the First and Second Doses, 12-17-Year-Olds, Quebec and British Columbia, Canada

| Interval between the 1st and 2nd doses | Quebec | | | | British Columbia | | | |
|---|---|---|---|---|---|---|---|---|
| | No. of cases | No. of controls | Unadjusted model VE (95% CI), % | Adjusted model [a] VE (95% CI), % | No. of cases | No. of controls | Unadjusted model VE (95% CI), % | Adjusted model [a] VE (95% CI), % |
| **14-27 DPV** | | | | | | | | |
| Unvaccinated | 1752 | 2140 | Reference | Reference | 665 | 944 | Reference | Reference |
| 21-34d (3-4wk) | 17 | 38 | 45.3 (2.8, 69.3) | 54.0 (16.8, 74.6) | 1 | 5 | NE | NE |
| 35-55d (5-7wk) | 9 | 38 | 71.1 (40.0, 86.0) | 75.6 (48.0, 88.6) | 3 | 8 | NE | NE |
| 56-83d (8-11wk) | 12 | 90 | 83.7 (70.2, 91.1) | 80.5 (63.6, 89.5) | 2 | 8 | NE | NE |
| 84+d (12+wk) | 7 | 58 | 85.3 (67.6, 93.3) | 86.0 (68.8, 93.7) | 3 | 11 | NE | NE |
| **28-55 DPV** | | | | | | | | |
| Unvaccinated | 1752 | 2140 | Reference | Reference | 665 | 944 | Reference | Reference |
| 21-34d (3-4wk) | 79 | 177 | 45.5 (28.4, 58.5) | 56.1 (41.6, 67.0) | 11 | 33 | 52.7 (5.7, 76.3) | 54.5 (7.9, 77.5) |
| 35-55d (5-7wk) | 36 | 113 | 61.1 (43.1, 73.4) | 64.2 (46.8, 76.0) | 9 | 48 | 73.4 (45.4, 87.0) | 69.0 (35.2, 85.2) |
| 56-83d (8-11wk) | 42 | 159 | 67.7 (54.4, 77.2) | 58.3 (39.9, 71.1) | 8 | 25 | 54.6 (-1.3, 79.6) | 57.9 (3.6, 81.6) |
| 84+d (12+wk) | 25 | 87 | 64.9 (45.0, 77.6) | 63.5 (42.0, 77.1) | 8 | 16 | NE | NE |
| **56-83 DPV** | | | | | | | | |
| Unvaccinated | 1752 | 2140 | Reference | Reference | 665 | 944 | Reference | Reference |
| 21-34d (3-4wk) | 223 | 375 | 27.4 (13.3, 39.2) | 42.7 (30.7, 52.6) | 39 | 59 | 6.1 (-42.4, 38.1) | 14.2 (-31.7, 44.1) |
| 35-55d (5-7wk) | 85 | 158 | 34.3 (13.8, 49.9) | 44.5 (26.2, 58.3) | 42 | 92 | 35.2 (5.4, 55.6) | 42.5 (14.9, 61.1) |
| 56-83d (8-11wk) | 50 | 160 | 61.8 (47.2, 72.4) | 45.0 (22.0, 61.2) | 21 | 45 | 33.8 (-12.3, 60.9) | 36.9 (-9.1, 63.5) |
| 84+d (12+wk) | 18 | 118 | 81.4 (69.3, 88.7) | 80.4 (67.1, 88.2) | 6 | 38 | 77.6 (46.7, 90.6) | 79.0 (49.3, 91.3) |
| **84-111 DPV** | | | | | | | | |
| Unvaccinated | 1752 | 2140 | Reference | Reference | 665 | 944 | Reference | Reference |
| 21-34d (3-4wk) | 526 | 874 | 26.5 (16.7, 35.1) | 43.5 (35.3, 50.6) | 63 | 117 | 23.6 (-5.5, 44.6) | 29.1 (1.1, 49.3) |
| 35-55d (5-7wk) | 103 | 216 | 46.7 (30.6, 59.1) | 46.3 (30.7, 58.4) | 75 | 126 | 15.5 (-14.4, 37.6) | 27.3 (0.5, 46.9) |
| 56-83d (8-11wk) | 45 | 152 | 63.8 (49.3, 74.2) | 60.6 (43.9, 72.3) | 22 | 63 | 50.4 (18.6, 69.8) | 50.3 (17.2, 70.2) |
| 84+d (12+wk) | 110 | 371 | 63.8 (54.8, 71.0) | 70.0 (62.1, 76.3) | 20 | 65 | 56.3 (27.2, 73.8) | 57.5 (28.0, 74.9) |
| **112-139 DPV** | | | | | | | | |
| Unvaccinated | 1752 | 2140 | Reference | Reference | 665 | 944 | Reference | Reference |
| 21-34d (3-4wk) | 446 | 717 | 24.0 (13.1, 33.5) | 34.3 (24.3, 43.0) | 67 | 148 | 35.7 (12.8, 52.6) | 42.2 (20.8, 57.8) |
| 35-55d (5-7wk) | 225 | 504 | 45.5 (35.4, 54.0) | 48.7 (38.4, 57.2) | 145 | 410 | 49.8 (37.8, 59.5) | 43.5 (28.8, 55.2) |
| 56-83d (8-11wk) | 2158 | 4373 | 39.7 (34.6, 44.4) | 50.6 (45.2, 55.4) | 129 | 494 | 62.9 (53.9, 70.2) | 57.7 (45.9, 67.0) |
| 84+d (12+wk) | 252 | 467 | 34.1 (22.2, 44.2) | 40.4 (28.8, 50.2) | 23 | 48 | 32.0 (-12.9, 59.0) | 38.4 (-4.0, 63.6) |
| **140-167 DPV** | | | | | | | | |
| Unvaccinated | 1752 | 2140 | Reference | Reference | 665 | 944 | Reference | Reference |
| 21-34d (3-4wk) | 371 | 752 | 39.7 (30.7, 47.6) | 39.5 (29.8, 47.8) | 54 | 84 | 8.7 (-30.3, 36.1) | 19.5 (-17.3, 44.8) |
| 35-55d (5-7wk) | 2977 | 5629 | 35.4 (30.2, 40.2) | 45.6 (40.2, 50.6) | 673 | 1352 | 29.3 (19.1, 38.3) | 33.3 (21.6, 43.2) |
| 56-83d (8-11wk) | 2471 | 4182 | 27.8 (21.8, 33.4) | 44.4 (38.9, 49.3) | 736 | 1349 | 22.6 (11.5, 32.3) | 32.2 (20.6, 42.2) |
| 84+d (12+wk) | 78 | 210 | 54.6 (40.7, 65.3) | 41.3 (21.4, 56.1) | 9 | 19 | NE | 49.9 (-15.5, 78.3) |

Abbreviation: d, days; DPV, days post-vaccination; NE, not estimable or 95% confidence interval spans ≥100%; VE, vaccine effectiveness; wk, weeks.
[a] Adjusted for age, sex, epi-week and region.

**Supplementary Table 6.** Profile of Participants (Adolescents Aged 12-17 Years), by Case/Control and Vaccine Status (Regardless of Time Since Vaccination), British Columbia, Canada, Epi-weeks 6-17

| | British Columbia | | | |
|---|---|---|---|---|
| | All (n=808) | | Three-dose vaccinated (n=432) | |
| **Characteristic** | **Cases, No. (column %) (n=144)** | **Controls, No. (column %) (n=664)** | **Cases, No. (row %) (n=42)** | **Controls, No. (row %) (n=390)** |
| **Age, years** | | | | |
| 12-14 | 72 (50.0) | 277 (41.7) | 15 (20.8) | 143 (51.6) |
| 15-17 | 72 (50.0) | 387 (58.3) | 27 (37.5) | 247 (63.8) |
| Median (IQR), years | 14.5 (13-16) | 15 (13-16) | 15 (14-16) | 15 (14-16) |
| **Sex** | | | | |
| Female | 79 (54.9) | 378 (56.9) | 29 (36.7) | 229 (60.6) |
| Male | 65 (45.1) | 286 (43.1) | 13 (20.0) | 161 (56.3) |
| **Infection severity** | | | | |
| Hospitalization [a] | 5 (3.5) | NA | 1 (20.0) | NA |
| **Epidemiological period** | | | | |
| 6-9 (Feb 2, 2022 – Mar 5, 2022) | 91 (63.2) | 248 (37.3) | 23 (25.3) | 134 (54.0) |
| 10-13 (Mar 6, 2022 – Apr 2, 2022) | 17 (11.8) | 195 (29.4) | 9 (52.9) | 117 (60.0) |
| 14-17 (Apr 3, 2022 – Apr 30, 2022) | 36 (25.0) | 221 (33.3 | 10 (27.8) | 139 (62.9) |
| **Time since third dose vaccination (days/weeks)** | **(all % values below this row are column %)** | | | |
| 0-13d (0-1wk) | NA | NA | 18 (42.9) | 102 (26.2) |
| 14-27d (2-3wk) | NA | NA | 8 (19.1) | 85 (21.8) |
| 28-55d (4-7wk) | NA | NA | 13 (31.0) | 115 (29.5) |
| 56-83d (8-11wk) | NA | NA | 3 (7.1) | 87 (22.3) |
| 84-111d (12-15wk) | NA | NA | 0 (0.0) | 1 (0.3) |
| Median (IQR), days | NA | NA | 17 (7-39) | 30 (13-54) |
| **Interval between 2nd and 3rd doses (days/weeks)** | | | | |
| 168-195d (24-27wk) | NA | NA | 11 (26.2) | 123 (31.5) |
| 196-223d (28-31wk) | NA | NA | 25 (59.5) | 241 (61.8) |
| 224-251d (32-35wk) | NA | NA | 6 (14.3) | 21 (5.4) |
| 252+d (36+wk) | NA | NA | 0 (0.0) | 5 (1.3) |
| Median (IQR), days | NA | NA | 202 (193-213) | 200 (193-208) |

Abbreviation: d, days; IQR, interquartile range; NA, not applicable; VE, vaccine effectiveness; wk, weeks.

[a] COVID-19 related hospitalization occurring within 30 days of the reference date.

**Supplementary Table 7.** Profile of Participants (Adolescents Aged 12-17 Years), by Case/Control and Vaccine Status (Regardless of Time Since Vaccination), Quebec, Canada, Epi-weeks 14-17

| Characteristic | Quebec | | | |
|---|---|---|---|---|
| | All (n=1012) | | Three-dose vaccinated (n=664) | |
| | Cases, No. (column %) (n=170) | Controls, No. (column %) (n=842) | Cases, No. (row %) (n=78) | Controls, No. (row %) (n=586) |
| **Age, years** | | | | |
| 12-14 | 78 (45.9) | 324 (38.5) | 17 (21.8) | 195 (60.2) |
| 15-17 | 92 (54.1) | 518 (61.5) | 61 (66.3) | 391 (75.5) |
| Median (IQR), years | 15 (13-16) | 15 (14-17) | 16 (15-17) | 15 (14-17) |
| **Sex** | | | | |
| Female | 91 (53.5) | 497 (59.0) | 47 (51.6) | 357 (71.8) |
| Male | 79 (46.5) | 345 (41.0) | 31 (39.2) | 229 (66.4) |
| **Infection severity** | | | | |
| Symptomatic | 60 (35.3) | NA | 27 (45.0) | NA |
| Hospitalization [a] | 5 (2.9) | NA | 1 (20.0) | NA |
| **Time since third dose vaccination (days/weeks)** | (all % values below this row are column %) | | | |
| 0-13d (0-1wk) | NA | NA | 5 (6.4) | 57 (9.7) |
| 14-27d (2-3wk) | NA | NA | 7 (9.0) | 68 (11.6) |
| 28-55d (4-7wk) | NA | NA | 37 (47.4) | 281 (48.0) |
| 56-83d (8-11wk) | NA | NA | 13 (16.7) | 111 (18.9) |
| 84-111d (12-15wk) | NA | NA | 13 (16.7) | 55 (9.4) |
| 112-139d (16-19wk) | NA | NA | 3 (3.8) | 12 (2.0) |
| 140-167d (20-23wk) | NA | NA | 0 (0.0) | 2 (0.3) |
| Median (IQR), days | NA | NA | 46.5 (38-73) | 44 (30-61) |
| **Interval between 2nd and 3rd doses (days/weeks)** | | | | |
| 168-195d (24-27wk) | NA | NA | 22 (28.2) | 112 (19.1) |
| 196-223d (28-31wk) | NA | NA | 32 (41.0) | 232 (39.6) |
| 224-251d (32-35wk) | NA | NA | 21 (26.9) | 184 (31.4) |
| 252+d (36+wk) | NA | NA | 3 (3.8) | 58 (9.9) |
| Median (IQR), days | NA | NA | 211.5 (193-227) | 218 (200-235) |

Abbreviation: d, days; IQR, interquartile range; NA, not applicable; VE, vaccine effectiveness; wk, weeks.

[a] COVID-19 related hospitalization occurring within 30 days of the reference date.

**Supplementary Table 8.** Unadjusted and Adjusted Three-Dose BNT162b2 Vaccine Effectiveness Against Omicron Infection (Epi-Weeks 6-17) by Vaccination Status, 12-17-Year-Olds, British Columbia, Canada

| | British Columbia | | | |
|---|---|---|---|---|
| | No. of | No. of | Unadjusted model | Adjusted model [a] |
| **Vaccination status** | **Cases** | **controls** | **VE (95% CI), %** | **VE (95% CI), %** |
| **Epi-weeks 6-17** | | | | |
| Unvaccinated | 102 | 274 | Reference | Reference |
| Three-dose vaccinated (0-13d) | 18 | 102 | 52.6 (17.8, 72.7) | 66.6 (39.0, 81.7) |
| Three-dose vaccinated (14+d) | 24 | 288 | 77.6 (64.0, 86.1) | 65.7 (41.9, 79.8) |

Abbreviation: d, days; VE, vaccine effectiveness.

[a] Adjusted for age, sex, epi-week and region.

**Supplementary Table 9.** Unadjusted and Adjusted Three-Dose BNT162b2 Vaccine Effectiveness Against Omicron Infection (Epi-Weeks 14-17) by Vaccination Status, 12-17-Year-Olds, Quebec, Canada

| | Quebec | | | |
|---|---|---|---|---|
| | | | Unadjusted model | Adjusted model [a] |
| **Vaccination status** | **No. of Cases** | **No. of controls** | **VE (95% CI), %** | **VE (95% CI), %** |
| **Epi-weeks 14-17** | | | | |
| Unvaccinated | 92 | 256 | Reference | Reference |
| Three-dose vaccinated (0-13d) | 5 | 57 | 75.6 (37.2, 90.5) | 76.5 (38.2, 91.1) |
| Three-dose vaccinated (14+d) | 73 | 529 | 61.6 (46.0, 72.7) | 63.3 (47.2, 74.6) |

Abbreviation: d, days; VE, vaccine effectiveness.

[a] Adjusted for age, sex, epi-week and region.
